# Parental Knowledge, Attitudes, and Practices Regarding Antibiotic Use in Pediatric Respiratory Infections: A Qualitative Study

**DOI:** 10.1101/2024.11.12.24317173

**Authors:** Muhammad Uneeb Khan, Zakir Khan, Areej Khan

## Abstract

Antibiotic misuse for respiratory tract infections (RTIs) is on the rise particularly among developing nations like Pakistan. This study evaluates parental knowledge, attitudes, and practices regarding RTI antibiotic use, which significantly impact antibiotic resistance trends. Targeted interventions to promote antibiotic use and reduce community resistance require understanding these dynamics.

Parents were recruited and interviewed in a tertiary care hospital and community pharmacies, representing both urban and rural areas, regardless of ethnicity or occupation. Face-to-face interviews with 21 parents were audio-recorded and transcribed verbatim. Conducted by one male and one female interviewer, these qualitative interviews provided an in-depth understanding of parental knowledge, attitudes, and practices (KAP) regarding antibiotic use for respiratory tract infections (RTIs) in children. Deductive thematic analysis was used, with predefined codes and themes refined throughout the process to capture evolving insights.

In total, 21 parents were interviewed, with 76% mothers and 24% fathers, and a median age of 24 for younger parents and 38 for older. Key barriers to accessing medical care for children with RTIs included financial constraints (38%) and transportation issues (34%). While 57% of parents preferred herbal remedies, 71% stated they would consult a doctor rather than reuse previously prescribed antibiotics. Additionally, 52% of parents believed doctors should take responsibility for educating families about proper antibiotic use and resistance.

This study highlights gaps in parental knowledge and practices regarding antibiotic use for RTIs in children, with financial and transportation barriers influencing access to medical care. While a significant portion of parents prefer herbal remedies, most would consult a doctor rather than reuse antibiotics. Targeted interventions, particularly through healthcare provider education, are crucial to promoting appropriate antibiotic use and combating resistance in developing nations like Pakistan.

## INTRODUCTION

Antibiotics saved millions of lives of people from fatal infections and allowed for surgical interventions and care of all age groups including infants and chemotherapy. The availability of antibiotics transformed healthcare from a diagnosis and prognosis-focused field to an interventional field which saved many lives daily. The evolution of resistance leading to the synthesis of new classes of antibiotics and misuse of new antibiotics led to further evolution of resistance and the cycle continued (Muteeb, Rehman, Shahwan, & Aatif, 2023). The CDC (Center of Disease Control) estimates that approximately 2.8 million people get infection from resistant bacteria in the United States, that are difficult to treat and result in 35000 deaths, annually (Sivasankar, Goldman, & Hoffman, 2023).

Antibiotic overuse may be exacerbated by a number of factors, including patients’ expectations, physicians’ expertise and experience (Albayrak, Karakaş, & Karahalil, 2021), pharmaceutical marketing, and the sale without prescription (Batista et al., 2020; Bhardwaj, Shenoy, Baliga, Unnikrishnan, & Baliga, 2021). Pakistan was ranked fifth among countries with the highest antimicrobial resistance (in 2019), further increasing during COVID-19 (Gul, Naveel, Khan, & Shah, 2023)

Respiratory infections range from mild conditions or non-significant illnesses such as the common cold to severe illnesses such as pneumonia.URTIs are more prevalent among children, particularly under six years of age. Children are more susceptible to developing respiratory tract infections due to a lack of immunity to viruses (Crawford & Davies, 2024).In Pakistan, 20-30% of deaths among children are attributed to acute RTIs (Naz et al., 2019). Children may be attributed to poor immunization rates and the wrong use of antibiotics to treat infections. Most of these respiratory infections are viral and can be tackled by preventive measures such as practicing excellent hygiene and washing hands frequently. (Toth, Rosenthal, Sharma, & Barnard, 2023).. However, Pakistan ranks as the third country with the most unvaccinated children around the globe (Saeed & Hashmi, 2021). Therefore, antibiotic misuse and abuse in Pakistan have led to a lower susceptibility and increased resistance to antibiotics that treat upper respiratory tract infections. (Khan et al., 2020; Mustafa et al., 2020)

Several studies evaluated that parents prefer self-medication without knowing the difference between bacterial and viral infections which leads to inappropriate use of antibiotics (Karatas et al., 2023; Sen Tunc, Aksoy, Arslan, & Kaya, 2021). The expectations of parents regarding the need for antibiotic prescription in acute viral infections have disrupted the rational prescribing pattern for respiratory infections. Further, the parents put pressure on the pediatricians to write antibiotics for their children on prescription orders (Grigoryan et al., 2006; Rousounidis et al., 2011). A study revealed that almost 72.6% of the attendants expect an antibiotic prescribed in RTIs (Abbasi & Nawaz, 2023).

Moreover, patients, doctors, and other healthcare professionals may misidentify bacteria from non-bacterial illnesses or misunderstand the effects of antibiotics, which leads to the inappropriate overuse of these drugs (Faiq).A study found that only 35.8% of antibiotics were appropriately prescribed according to the indication in RTIs among children of the Punjab (Ul Mustafa et al., 2024). From the perspective of healthcare providers, various factors such as limited understanding of Upper Respiratory Tract Infections (URTIs) and their appropriate management, erosion of trust between patients and providers, absence of clear prescription protocols and guidelines, inadequate stewardship of diagnostic and antibiotic prescriptions, insufficient utilization of evidence-based criteria for diagnosis and treatment, lack of expertise in the treatment and management of infectious diseases for pediatric patients, poor parent-provider communication, as well as sociodemographic and healthcare facility characteristics of providers, may contribute to the unnecessary prescribing of antibiotics for URTIs in children (Del Fiol et al., 2022; Pierantoni et al., 2021).

A comprehensive understanding of parental KAPs regarding antibiotic use in RTIs is necessary to progress on the national action plan (NAP) against AMR and achieve sustainable developmental goals (SDGs). Pakistan currently faces challenges in progressing with the national action plan on antimicrobial resistance (Ullah et al., 2022). Further, Sustainable Developmental Goal 3.2 (SDG 3.2) aims to eradicate childhood mortality due to acute RTIs such as pneumonia from Punjab by 2030 (Chaudhry & Khan, 2020; Organization, 2015).

In this study, we aim to evaluate the knowledge, attitudes, and practices of parents regarding the use of antibiotics for Respiratory tract infections in children. This study will not only allow an assessment of the progress of the national action plan for combating AMR in RTIs, but it will guide the development of targeted educational interventions and strategies to achieve SDG 3.2 i.e., to reduce childhood morbidity and mortality.

## METHODOLOGY

### Methods

For the Selection of parents to be interviewed, a convenience sampling strategy was employed and parents were interviewed until saturation point was met. Parents presented at tertiary care hospital and community pharmacies, were included in the study coming from both, urban and rural, practices. Parents were interviewed regardless of their ethnicity and occupation.

### Interviews

Two interviewers (one female and one male) conducted face to face interviews of parents presented at primary care. Interviews were audio-recorded and transcribed verbatim. Qualitative interviews were the most effective approach for obtaining in-depth understanding of parents’ knowledge, attitudes, and behaviors regarding antibiotic usage in respiratory tract infections (RTIs).

### Sample Size

An estimated sample size was 20 to 30 participants. The final sample size determined after reaching data saturation was 21.

### Analysis

Deductive thematic analysis was done in order to obtain views of parents KAPs and concerns about antibiotic use in RTI’s in children. Themes for the interviews were already defined with certain codes. After some initial interviews, themes and codes were refined during all this as this process entailed the connection, categorization, repositioning, renaming, addition, and removal of themes as necessary to generate a cohesive set of themes.

### Ethical Statement

In this study, data was collected through a questionnaire to evaluate parental knowledge, attitudes, and practices regarding antibiotic use for respiratory infections in children. The study did not involve any intervention, human specimens, tissues, or animal research. Ethical approval was obtained from REC(Riphah Ethical Committee) with approval number **REC/RIPS/2024/10** and informed consent was obtained from all participants prior to their participation in the survey

### Human Ethics and Consent of participation

Human ethics consent was not applicable to this research but consent to participation was applicable and consent was taken before data collection. Consent to participate statement is mentioned in Questionnaire.

### Clinical trial number

Clinical trail number : not applicable

## RESULTS

### Demographics

In total, 21 participants took part in this study in which 76% (n=16) mothers and 24% (n=5) fathers were interviewed.Nevertheless, any individual who assumed caring role could have taken part.71% (n=15) of mothers were House Wives,one mother and Five Fathers were employed. Median age of parents aging (18-30) is 24, and median age for parents aging (31-45) is 38 with average number of children 2 and only one parent was aging above 45.

**Table 1.** Demographics of the participants.

|  | Age | Gender | No Of Children |
| --- | --- | --- | --- |
| KAP-1 | 18-30 | F | 2 |
| KAP-2 | 31-45 | F | 3 |
| Kap-3 | 18-30 | F | 1 |
| KAP-4 | 31-45 | F | 2 |
| KAP-5 | 18-30 | F | 2 |
| KAP-6 | 18-30 | F | 1 |
| KAP-7 | 31-45 | F | 4 |
| Kap-8 | 18-30 | F | 3 |
| KAP-9 | 18-30 | F | 1 |
| KAP-10 | 18-30 | F | 2 |
| KAP-11 | 31-45 | F | 3 |
| KAP-12 | 18-30 | F | 1 |
| KAP-13 | 31-45 | F | 4 |
| KAP-14 | 31-45 | F | 1 |
| KAP-15 | 18-30 | F | 2 |
| KAP-16 | 31-45 | M | 2 |
| KAP-17 | 31-45 | M | 2 |
| KAP-18 | >45 | M | 3 |
| KAP-19 | 31-45 | M | 1 |
| KAP-20 | 31-45 | F | 1 |
| KAP-21 | 31-45 | M | 3 |

### Results of phase 2

By Deductive thematic analysis,we had defined themes but we identified few sub-themes in our data regarding parents concerns and practices about antibiotic use in children with RTI’s. Themes and Sub-themes are described in the following section.

#### 1. Barriers while acquiring medical facilities

Pakistan being a third world country has a lot of loopholes in their medical sector so considering this parents were asked about barriers they face when their child has RTI and they have to acquire medical facilities to treat their child.

##### a) Financial barriers

As Pakistan’s inflation rate is currently at rise, it has become difficult for people to afford and avail medical facilities where 29% (n=6) of parents are earning (31k-50k) with median income 40.5k per month, 52% (n=11) earning (51k-100k) with median income 75.5k per month and 19% (n=4) earning above 100k per month.(Incomes are in Pakistani rupees) It has become difficult to manage household with expensive medicine and expensive lab tests and without lab reports it is difficult for physician to diagnose properly and high chances of being treated wrongly. 38% (n=8) parents stated that they face financial issues while acquiring medical facilities for their children with RTI.

##### b) Travelling barriers

Pakistan being under-developed country has no proper transport system and people face a lot of transportation issues. Pakistan also face traditional and cultural issues as most of mother parents are not allowed to go for checkup without father and adding to all these factors, financial burden is real, which makes it more difficult to travel. 34% (n=7) parents mentioned travelling and transportation a barrier with multiple reasons i.e no hospital nearby and if they have facility of hospital nearby,availability of doctor becomes issue which makes it difficult for parents to avail medical facilities for their children with RTI.

##### c) Appointment barrier

Taking appointment for checkup becomes difficult sometimes but most of the parents were satisfied that they never faced appointment issues. Only one parent mentioned that they find this as a barrier when they are unable to get appointment for their child.

##### d) Finding a good docto**r**

To some parents it was difficult to find and choose a specialist doctor for their child. One of the parent stated this as

> *“first of all finding a good doctor is one of the problems because doctors these days put kids on antibiotics or high potency drugs right away so you to have to make sure you find a doctor who is not prescribing kids antibiotics or high potency drugs straight away**’** (KAP-1,children-2)*

#### 2. Role of Lifestyle

For Children being prone to RTI’s, their lifestyle matters a lot. All of the parents agreed that diet and hygiene are the key factors that can save their children staying away from catching RTI’s. Few quotes are illustrated below,

> “*I make sure they wear a mask and wash their hands frequently. I also make them eat and drink in separate utensils and they are not allowed to have any sour or cold beverage.” (KAP-7,Children-4)*

> “ *it has a great role in it actually because these diseases only spread because unhygienic conditions. If kid’s hands are dirty or he is drinking from a dirty bowl or glass then it can spread from those things. If a kid is being provided with a clean environment then I think that kid can be protected from the diseases.” (KAP-1, Children-2)*

> “*First of all cleanliness is the most important factor because little things like dust or unhygienic living environments play a major role in respiratory tract infections*. *Parents should make sure their children are not consuming any oily or spicy food because such eatables can cause infections in chest or disrupt the process of respiration.*” *(KAP-16, children-2)*

#### 3. Alternative Therapy

In this section, we found that antibiotics are not the sole solution for treating RTI’s. Parents talked about using herbal remedies for treatment of their children.33% (n=7) parents said that they do not use any herbal remedy for treating their children.

10%(n=2) of the parents said that they once used herbal remedies but found them ineffective so they will not use them in future.

> *“I used to go for herbal remedies but they’re on the bottom of my to-do list as being a working lady it is hard to keep up with regularity and herbal remedies demand consistency. Meanwhile antibiotics are quick in terms of effectiveness so now I prefer antibiotics.”*(*KAP-6, Children-1)*

> “*For once or twice have I used herbal tea for cough which an elderly person of my family told me but I did not see any positive results. Medicines works fine for my kids” (KAP-2,children-3)*

And then there were 57% (n=12) of the parents who stated that they use and prefer Herbal remedies for the treatment of RTI’s. Most used herbal remedies were *Cinnamon Tea, Clove Tea, Garlic Tea and Honey mixed with Ginger Juice.* Parents while telling about herbal remedies mentioned that these remedies are very effective. Few quotations are illustrated below;

> “*I do use herbal remedies for cough. The elderly women in my family recommend honey for treating cough in children. Sometimes we drink soaked local herbs with boiled water to treat severe cough.” (KAP-5, children-2)*

> “*I prefer herbal remedies over antibiotics. Tea without milk is preferred in my family and steam is also a good option.” (KAP-7, children-4)*

> “*I do use herbal remedies. I mostly use ‘Clove Tea’ for everyday cold. I don’t really believe in antibiotics because they have more side effects than benefits. Actually, I’m from a backward area so it’s nearly impossible for me to get a medicine on time. We sometimes use ‘Joshanda’ as well but mostly clove tea.*” (*KAP-20, Children-1*)

#### 4. Impact of Past Experiences

If a child had an RTI and it was treated with antibiotics, what will be Parent’s attitudes towards RTIs in future on the basis of past experiences, were assessed in this section. Parents talked about the impact of past experience and it was determined that 71% (n=15) parents will consult to doctor rather then re-using the same medication again. Some of the quotations are illustrted below;

> “*If my kid has suffered from any disease or condition in the past and all of the sudden he/she faces it again I won’t reuse the previously prescribed medicine. I think no parents should do that*” (*KAP-21, Children-3)*

> *“If my child faces any issue which she have suffered from in past, I’ll take her to the doctor. I won’t reuse the previously prescribed medicines.” (KAP-5, children-2)*

Whereas, 29% (n=6) were of this view that they can re-use the same medicine again rather then consulting doctor. Some quotations are illustrated below;

> “*My child have never suffered from any chronic medical issue in the past. It just the fever and common cold so initially I’ll prefer the same previously prescribed medicine if conditions remains the same then I’ll visit the doctor.” (KAP-6, children-1)*

> “*First I’ll try to treat my kid by home remedies and its 70% effective then go to medicines that are needed to recover completely.” (KAP-7, children-4)*

> “*First we’ll try to treat our kids at home by reusing the previously prescribed medicines.*” *(KAP-17, children-2)*

This re-use of previously prescribed medicine may also fall under category of Self-medication.

These attitude and beliefs of Self medicating and Re-using the same medicine can be linked with below mentioned factors;

##### a) Educational Backgrounds

Out of those 6 parents who opted to re-use the medicine 67% (n=4) are less educated (KAP-7/College, KAP-12/ Uneducated, KAP-17/Primary school, KAP-20/Secondary school).

##### b) Residence

Out of those 6 parents who opted to re-use the medicine 67% (n=4) are residents of rural areas.(KAP-7,KAP-12, KAP-17, KAP-20-Rural areas).

These above mentioned factors cannot be spared as they are playing major role in such practices.

#### 5. Suggestions about awareness of antibiotic use and antibiotic resistance

It is very important to address people about proper use of antibiotics and also awareness to be given about Antibiotic resistance as misuse and abuse will lead to ineffectiveness of antibiotics which may result in disaster for the community.

38% (n=8) parents were of this view that Social media campaigns should be run to spread awareness in common people as mostly everyone uses social media, so this can be an effective and quicker method to spread awareness.10% (n=2) stated that Seminars should be conducted for spreading awareness. 52% (n=11) parents said that it should be Doctor’s duty to counsel about proper use of antibiotics as everyone does not know this. Some quotations are illustrated below;

> “*There should be seminars regarding antibiotic resistance specifically for mothers. Some mothers use the same medicine again and again and which ultimately pose negative impacts on their child’s health. “ (KAP-3, children-1)*

> *“I think should organize campaigns on social media because everyone has now access to internet. And it is not possible for everyone to attend such workshops physically.” (KAP-6, Children-1)*

> *“ Doctors should perform experiments on dummies to portray the phenomenon of antibiotic resistance. Distribution of pamphlets with all basic knowledge can also be a good option to make people aware of antibiotic resistance.” (KAP-9, children-1)*

> *“ Social Media should be used to run campaigns. Facebook is quite popular in this regard.” (KAP-11, children-3)*

### Major Findings

The analysis of parental apprehensions and methodologies concerning the administration of antibiotics to children with respiratory tract infections (RTIs) revealed various significant themes and sub-themes. Parents encountered significant obstacles in accessing medical facilities, primarily due to financial, travel, and appointment barriers. Among these, financial constraints were the most prominent, exacerbated by the increasing inflation. Parents also emphasized the challenge of locating a competent physician who refrains from promptly prescribing potent antibiotics. The significance of lifestyle factors, specifically diet and hygiene, in the prevention of respiratory tract infections (RTIs) was acknowledged. Numerous parents emphasized the necessity of upholding cleanliness and adhering to a healthy diet. The majority of parents preferred alternative therapies, such as herbal remedies, with cinnamon and clove tea being the most popular. However, some parents still favored antibiotics due to their faster efficacy. Prior encounters with RTIs had an impact on subsequent attitudes, as the majority of parents chose to seek medical advice rather than reusing previous medications. However, a small percentage, especially those with lower educational backgrounds or residing in rural areas, were inclined towards self-administration of medication. Finally, there was an agreement on the necessity of increasing awareness regarding appropriate antibiotic usage and resistance. Proposed strategies to tackle this issue effectively include social media campaigns, seminars, and counseling led by doctors.

## DISCUSSION

The misuse of antibiotics in respiratory tract infections among children can lead to the development of antimicrobial-resistant bacteria. Parents are often responsible for the healthcare needs of their children and their knowledge and practices can significantly impact the use of antibiotics in their children. Therefore, this study assessed the parent’s knowledge, attitudes, and practices regarding antibiotic use among children with respiratory tract infections by interviews to generate a holistic overview of the matter.

Most of the parents included in this study had one to two children and a significant proportion frequently experienced chronic respiratory disease. We found that most of the parents had poor knowledge about antibiotics and many misconceptions regarding the treatment of respiratory tract infections with antibiotics. Further, they had a positive attitude but poor practices regarding antibiotic use in children.

Most of the parents had poor knowledge regarding antibiotics and believed antibiotics to be the primary treatment against respiratory tract infections, similar to other studies conducted in Saudi Arabia, Greece, and Ethiopia respectively (Alzaid et al., 2020; Oikonomou et al., 2021; Zeru, Berihu, Buruh, Gebrehiwot, & Zeru, 2020). Previous research shows that parents with good knowledge of antibiotics do not misuse them in acute viral respiratory infections and vice versa (Almohideb, 2020; Zeru et al., 2020). Moreover, studies show that improper use of antibiotics in viral infections can kill susceptible bacteria, disturb natural flora, and allow susceptible bacteria to survive and multiply (Godman et al., 2020; Tang, Millar, & Moore, 2023). This leads to death from common infections that could be treated otherwise.

Additionally, the misuse of antibiotics in viral infections has more harm and financial loss than benefits. For instance, a study assessed the impact of antibiotics on treatment outcomes in hospitalized patients with active respiratory infections and found that late initiation of antibiotics increased mortality but had no positive effect on the outcome, whether antibiotics were given or not (Hovind, Berdal, Dalgard, & Lyngbakken, 2024). However, the in-hospital stay and cost increased with antibiotic therapy in respiratory infections (Hovind et al., 2024). Therefore, poor parental knowledge may lead to an increase in AMR and healthcare costs.

Moreover, parental misconceptions such as believing that antibiotics are effective against viral infections, and do not produce any side effects can lead to increased antibiotic misuse and overuse, consequently exacerbating the issue of antimicrobial resistance. A systematic review emphasized that even parents with good knowledge regarding the use of antibiotics often misuse them solely due to misconceptions similar to the ones found in our study (Cantarero-Arévalo, Hallas, & Kaae, 2017). Additionally, the lack of awareness regarding the potential side effects of antibiotics is one of the contributing factors leading to parents’ attitudes toward insisting physicians for prescribing antibiotics (Biezen, Grando, Mazza, & Brijnath, 2019). Other studies show that several cultural factors as well as parent’s previous experiences with antibiotic treatments shape their demand for antibiotics (Ledingham, Hinchliffe, Jackson, Thomas, & Tomson, 2019).

The parents reported in interviews that they prefer a physician who does not prescribe antibiotics in the first instance. Research studies in other regions also found a similar gap in parental knowledge and practices where parent’s knowledge regarding antibiotics was not in line with their practices regarding antibiotic use in viral infections (Harun, Haider, Haque, Chowdhury, & Islam, 2020; Paredes et al., 2022). The majority of parents had poor practices regarding antibiotic use, but they did not prefer to self-medicate with the same antibiotics as for previous infections. Instead, they tended to traditional herbal remedies. While there exists moderate evidence for the effectiveness of herbal remedies in symptomatic relief and antibacterial effects (Anheyer, Cramer, Lauche, Saha, & Dobos, 2018). There is a lack of evidence on the effectiveness of these remedies in treating RTIs. Additionally, in the case of bacterial RTIs, these remedies alongside the practice of self-medication may lead to delays in seeking physician assessments and thereby higher number of complications (Martin & Ernst, 2003).

Poor practices were more prevalent among parents whose children had chronic respiratory infections similar to the findings of Zyoud et al. (2015). This may be attributed to the frequent use of antibiotics in children with chronic infections that make parents believe that antibiotics are necessary for treatment leading to pressure on healthcare providers and self-medications. Moreover, parental practices regarding antibiotic use were influenced by their level of education irrespective of their knowledge and attitudes. Therefore, educational interventions focusing on improving parental knowledge and behaviors may lead to improved practices and a lesser risk of AMR.

Furthermore, parents reported a lack of accessibility and affordability to seek timely treatment from healthcare providers. In most low and middle-income countries, the healthcare services are geographically distant and the healthcare services are expensive while the families have limited financial resources (Dawkins et al., 2021). These conditions lead to delays in treatments and consequently increased complications (Dawkins et al., 2021). Similarly, cultural barriers were found to hinder timely treatments due to cultural barriers such as the necessary male companions to travel even towards healthcare setups. These cultural barriers can be addressed by promoting awareness through community leaders and community health workers (Sarin & Lunsford, 2017). Parents agreed to the need for awareness regarding antibiotic use and resistance and most of them trusted that doctors could play a significant role in counseling patients. Some of them also suggested social media campaigns and seminars.

### Conclusion

The study highlights the significant role that parental knowledge, attitudes, and practices play in the use of antibiotics for respiratory tract infections (RTIs) in children. Financial and logistical barriers were identified as major obstacles to accessing proper medical care, while cultural preferences, such as the use of herbal remedies, also influenced treatment choices. Although the majority of parents preferred seeking medical advice over reusing antibiotics, a notable proportion practiced self-medication, often linked to lower education levels and rural residency. The findings underscore the need for targeted interventions, including social media campaigns and physician-led counseling, to address antibiotic misuse and raise awareness about antibiotic resistance.

## Data Availability

Already provided as part of submitted article

